# Causal effects from tobacco smoking on obesity-related traits: a Mendelian randomization study

**DOI:** 10.1101/2022.06.27.22276929

**Authors:** Sehoon Park, Seong Geun Kim, Soojin Lee, Yaerim Kim, Semin Cho, Kwangsoo Kim, Yong Chul Kim, Seung Seok Han, Hajeong Lee, Jung Pyo Lee, Kwon Wook Joo, Chun Soo Lim, Yon Su Kim, Dong Ki Kim

## Abstract

**Background:** There is a notion that tobacco smoking would have weight control effect based on the appetite suppressive effect of nicotine. However, a causal effect from being an ever smoker on obesity-related traits in the general population has yet been determined.

**Methods:** This Mendelian randomization (MR) analysis instrumented 378 genetic variants associated with being an ever smoker which mostly initiated in adolescents or young adulthood, identified from a genome-wide association study (GWAS) meta-analysis of 1.2 million individuals. The outcome data for body mass index, waist circumference, hip circumference, and waist-to-hip ratio was collected in 337,318 white British ancestry UK Biobank participants with 40-69 ages. Replication analysis was performed for GWAS meta-analysis for body mass index including the GERA/GIANT data including 364,487 mostly European samples. Summary-level MR by inverse variance weighted method and pleiotropy-robust MR methods, including median-based and MR–Egger regression, was performed.

**Results:** Summary-level MR analysis indicated that genetically predicted being an ever smoker is causally linked to higher body mass index [+0.28 (0.18, 0.38) kg/m^2^], waist circumference [+0.88 (0.66, 1.10) cm], hip circumference [+0.40 (0.23, 0.57) cm], and waist-to-hip ratio [+0.006 (0.005, 0.007)]. The results were consistently supported by pleiotropy-robust MR analysis. In the replication analysis, genetically predicted being an ever smoker was again significantly associated with higher body mass index [+0.03 (0.01, 0.05) kg/m^2^].

**Conclusion:** Initiation of tobacco use may consequently lead to worse obesity-related traits of the general population in middle-to-old ages.

## Background

Tobacco smoking is an important global health problem which increases risks of cardiovascular disease, malignancy, and death.^1^ Tobacco smoking is responsible for more than 8 million deaths a year, and nearly 1.2 million individuals die related to second-hand smoking. Medical society pays large efforts to reduce tobacco use, and some promising results suggests the socioeconomic burden related to smoking might have started to decrease. There has been a notion that tobacco smoking reduces weight.^2^ The concept of slimness related to tobacco use has been even advertised by tobacco companies and the misconception considering tobacco as weight control measure led young adolescents to initiate smoking.^3,4^ That nicotine suppresses appetite in short-term and weight gain phenomenon after smoking cessation even consolidated the misbelief that smoking would be helpful to reduce weight.^5,6^ On the other hand, recent studies demonstrated that being obese causes smoking initiation, suggesting the causal linkage between smoking and obesity.^7^ Yet, the causal effects from smoking initiation on obesity-related traits in the general population warrants additional study. A population-scale evidence testing whether smoking initiation truly leads to lower weight in middle-to-old age individuals would be important for public policy. Furthermore, if the notion turns out to be false, the information should be widely used for social education to prevent tobacco initiation in the vulnerable population who may use tobacco anticipating the weight control effect.^3^

Mendelian randomization (MR) is an analytic tool to investigate causal effects implementing genetic instrumental variable.^8^ MR has been widely used in medical literature testing the causal estimates from modifiable exposure on health outcomes. As genetic instrumental variable is fixed before birth, MR can demonstrate causal estimate minimally biased from confounding or reversal causal effects.

In this study, we aimed to test the causal effects from tobacco smoking on obesity-related traits. We performed MR analysis in two-large scale genetic databases. We found that being an ever smoker may consequently leads to higher body mass index and worse obesity-related characteristics in middle-to-old age populations.

## Methods

### Ethical considerations

The study was approved by the institutional review boards of Seoul National University Hospital (No. E-2203-053-1303) and the UK Biobank consortium (application No. 53799).^9,10^ The study was performed in accordance with the Declaration of Helsinki. The requirement for informed consent was waived by the attending institutional review boards because public anonymous database collected for research purpose was used for this study.

### Genetic instruments for tobacco smoking

We used the findings from one of the largest GWAS meta-analysis for over 1.2 million European ancestry individuals including over half a million ever smokers (Figure 1).^11^ In the meta-analysis, 29 studies were included and the representative studies which provided relatively large number of samples were 23andMe, UK Biobank, deCODE, HUNT and others. Total of 378 lead single nucleotide polymorphisms (SNPs) with conditionally genome-wide significant association with being an ever smoker was used as the genetic instrumental variable in this study (Supplementary Table 1). The mean age of tobacco smoking initiation ranged from 13.7 to 22.9 years old in the dataset, indicating the study analyzed being an ever smoker which mostly initiated in adolescents or in early periods of adulthood as in the general population.

**Figure 1.**
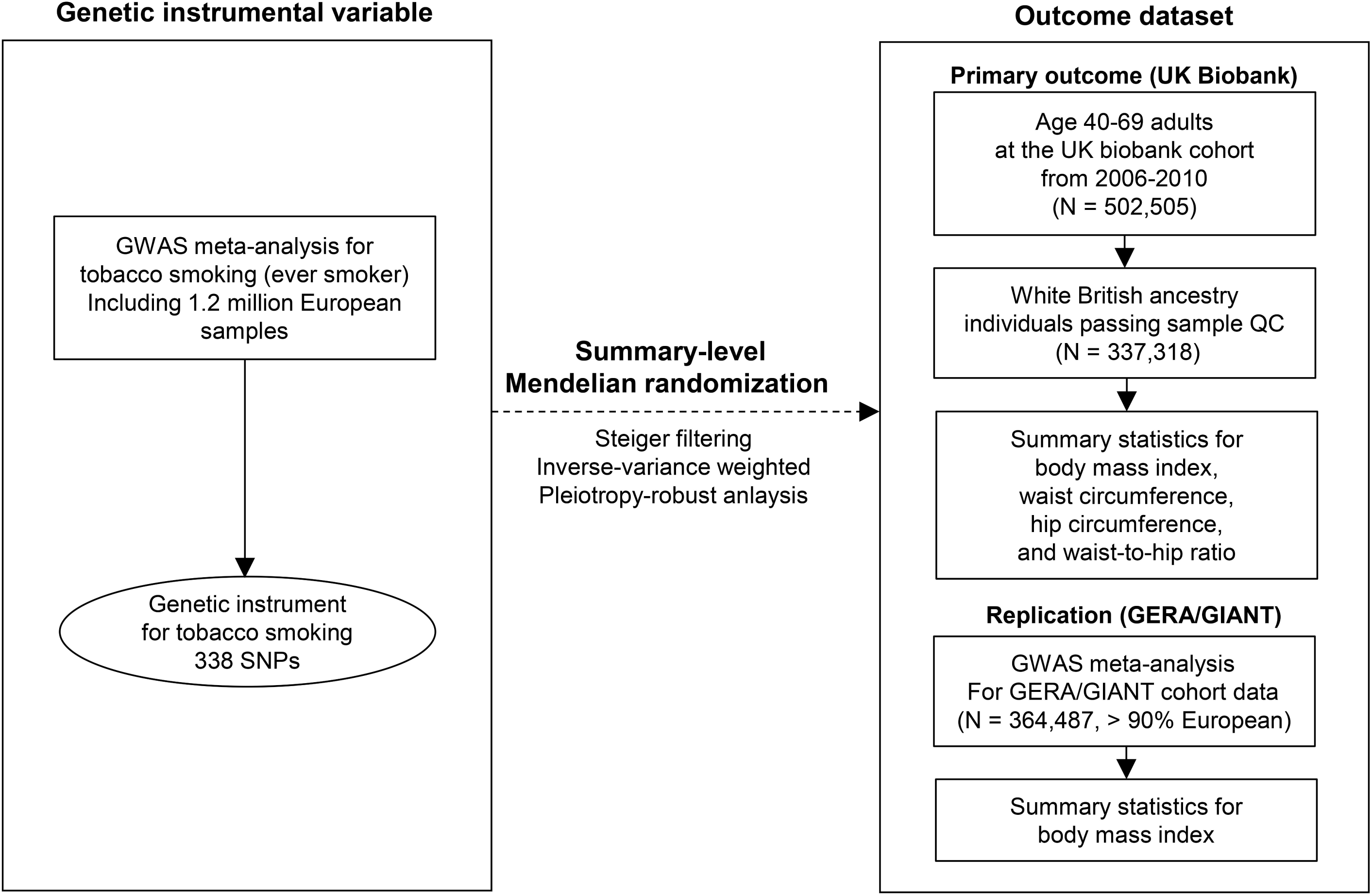
Study flow diagram. GWAS = genome-wide association study, QC = quality control, SNPs = single nucleotide polymorphism

### MR assumptions

A valid MR analysis should attain three core assumptions to demonstrate causal effects.^8^ First, the relevance assumption is that the genetic instrumental variable should be strongly associated with the exposure of interest. We collected genetic variants with genome-wide significant association with the tobacco smoking phenotype, explaining ∼2% variance for smoking initiation,^11^ thus, the assumption was considered attained. Second, the independence assumption is that the instrumental variable should not be associated with a confounder. We performed pleiotropy-robust MR analysis which can relax the assumption and statistically tested the possibility of a directional pleiotropy (e.g. MR-Egger regression).^12^ Also, we performed sensitivity analysis with the scanned results using the GWAS catalog to screen any potentially confounder-associated SNPs.^11,13^ We performed the sensitivity analysis after excluding the SNPs with identified significant association with other traits. Third, the exclusion-restriction assumption is that the tested causal effect should be through the exposure of interest. Although this assumption is not statistically testable, we performed MR analysis by median-based methods which relaxes the assumption for up to half of the instruments, serving as a sensitivity analysis.^14^

### Steiger filtering

That obesity causally affects tobacco smoking behavior has been reported previously.^7^ In such situation, reverse directional genetic effect should be carefully considered for an MR analysis. Namely, a genetic variant that has stronger association with an outcome phenotype than with an exposure may reflect the reverse genetic effect from outcome to exposure, thus, should be removed from the genetic instrumental variable. Steiger filtering provides the statistical method to test the direction of the effect from a genetic variant, comparing the explained variance for exposure and for outcome phenotypes.^15^ We used the process with the UK Biobank data and the SNPs reflecting genetic effects with direction from tobacco smoking on obesity-related traits were implemented in this study.

### UK Biobank outcome dataset and phenotypes

The UK Biobank data was implemented as the main outcome dataset for this study (Figure 1).^9,10^ The main strength of the data is availability of various anthropometric parameters related to obesity and the large-scale genetic information for robustly quality controlled samples. The limitation of the dataset for this study is that the developmental GWAS meta-analysis for the genetic instrumental variable includes the UK Biobank data, and such sample overlap may cause bias towards observational confounding when instrumental strength is weak.^16^

Among the > 500,000 participants, we collected the information from 337,318 white British ancestry samples who passed sample quality control, and those who were outliers regarding heterogeneity, missing rate, with excess kinship, or sex chromosomal aneuploidy were excluded.^17,18^

We collected body mass index, waist circumference, hip circumference and waist-to-hip ratio which were measured at the baseline visits when the participants had age from 40 to 69. To calculate the association between genetic instruments and outcome phenotype, linear regression analysis was performed for the SNPs by PLINK 2.0 and the genetic effect sizes were adjusted for age, sex, age×sex, age^2^, and 10 genetic principal components.^19^

### Replication outcome dataset

In consideration for the sample overlap between the GWAS meta-analysis for tobacco smoking and the UK Biobank data, an independent replication outcome data was warranted. Two-sample MR with lower proportion of sample overlap has strength in that the analysis is less likely to be biased towards observational associations.^16^

The previous GWAS meta-analysis for body mass index using the Genetic Epidemiology Research on Adult Health and Aging (GERA) and Genetic Investigation of ANthropometric Traits (GIANT) consortium was used as the replication outcome data, which was available in the GWAS catalog (URL: https://www.ebi.ac.uk/gwas/studies/GCST006368) (Figure 1).^13,20^ The meta-analysis included 364,487 samples mostly of European ancestry towards body mass index phenotype (Supplementary Table 2). The GERA data was not included in the previous meta-analysis which provided the genetic instrumental variable, as GERA data was only used for GWAS towards drinking behavior in the meta-analysis.^11,21^ The GIANT data had some sample overlap (e.g. ARIC, EGCUT, HRS, METSIM, QIMR studies) with the GWAS meta-analysis for tobacco smoking,^22^ yet the number of samples included in the both dataset was ∼50,000 at maximum, which is much lower than the UK Biobank data has.

Regarding other obesity-related phenotypes, a summary statistic with large-scale sample size or sufficient number of genotyped SNPs was scarce, thus, we performed the replication analysis towards body mass index, which would be the primary parameter that defines obesity.

### Summary-level MR analysis

Among the 378 SNPs instruments, only a SNP (rs3076896) was not available in the UK Biobank data which was not included in the according analysis. However, larger number of SNPs were unavailable in the GERA/GIANT GWAS meta-analysis for body mass index, thus, we used proxy SNPs (linkage disequilibrium R2 > 0.8) to replace the missing. This process allowed us to include all SNPs except two (rs3076896, rs748832) among the variants passing the Steiger filtering process in the UK Biobank for body mass index trait. Next, the summary-level datasets were harmonized and the palindromic SNPs with intermediate allele frequencies were disregarded because of ambiguous strandness.

Multiplicative random-effect inverse variance weighted method was used to calculate main causal estimates following the current guideline for MR. Additional sensitivity analysis was performed including weighted median and simple median methods, which yields valid causal estimates even up to half of the instruments are invalid. The weighted median method allows invalidity in up to half of the instrumented weights, while the simple median method in up to half of the variants, having distinct strengths as pleiotropy-robust MR sensitivity analysis. MR-Egger regression with bootstrapped standard error was performed which relaxes the independence assumption under attainment of the InSIDE assumption. MR-Egger intercept P value was calculated which statistically tests the possibility of directional pleiotropy, and a significantly non-zero intercept (MR-Egger intercept P value < 0.05) was considered to indicate a possible pleiotropic effect. Bonferroni-adjusted two-sided P value of < 0.05/5 by the inverse variance-weighted method was used to indicate statistical significance. The causal estimates were scaled to the effects from two-fold increase in prevalence of being a smoker by multiplying 0.693 to the raw causal estimates.^23^ The summary-level MR analysis was performed by “TwoSampleMR” package in R (version 3.6.0) and the MR-base web platform.^24^

## Results

### Characteristics of the outcome datasets

The final UK Biobank outcome data had median age of 58 years old (interquartile range 51, 63) (Table 1). Male sex was identified for 46.3%, median body mass index was 26.7 (interquartile range 24.1; 29.9) kg/m^2^. The median values for waist circumference, hip circumference, and waist-to-hip ratio were 90 (80, 99) cm, 102 (97, 108) cm, and 0.874 (0.802, 0.937), respectively. In the data, the prevalence of diabetes mellitus was 4.8% and total of 20.9% and 17.5% of the samples were taking hypertension and dyslipidemia medications, respectively.

**Table 1.**
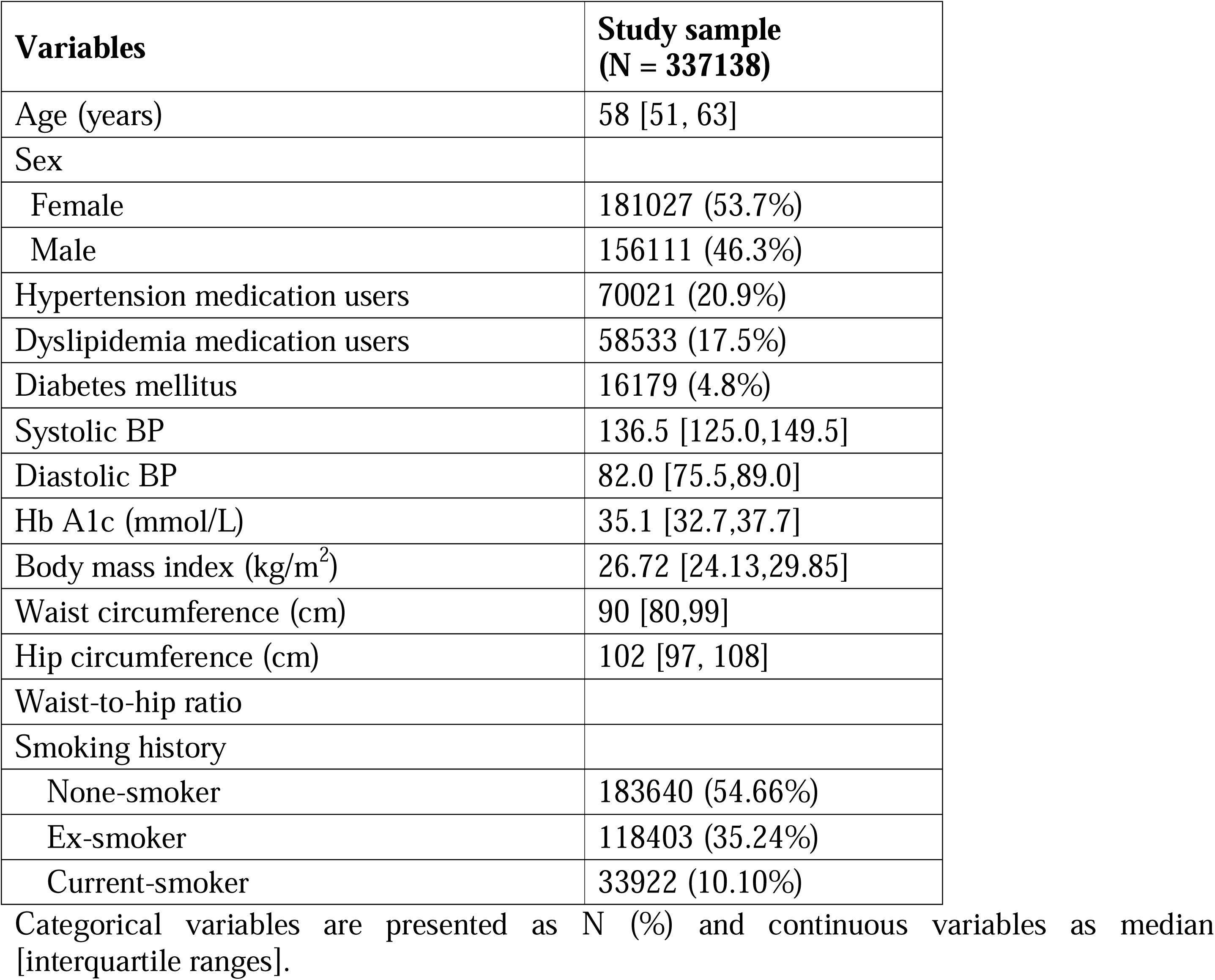
Characteristics of the UK Biobank data.

| **Variables** | **Study sample**<br>**(N = 337138)** |
| --- | --- |
| Age (years) | 58 [51, 63] |
| Sex |  |
| Female | 181027 (53.7%) |
| Male | 156111 (46.3%) |
| Hypertension medication users | 70021 (20.9%) |
| Dyslipidemia medication users | 58533 (17.5%) |
| Diabetes mellitus | 16179 (4.8%) |
| Systolic BP | 136.5 [125.0,149.5] |
| Diastolic BP | 82.0 [75.5,89.0] |
| Hb A1c (mmol/L) | 35.1 [32.7,37.7] |
| Body mass index (kg/m<sup>2</sup>) | 26.72 [24.13,29.85] |
| Waist circumference (cm) | 90 [80,99] |
| Hip circumference (cm) | 102 [97, 108] |
| Waist-to-hip ratio |  |
| Smoking history |  |
| None-smoker | 183640 (54.66%) |
| Ex-smoker | 118403 (35.24%) |
| Current-smoker | 33922 (10.10%) |

Categorical variables are presented as N (%) and continuous variables as median [interquartile ranges].

The GERA GWAS had mean ages of ∼60 years old including ∼ 60% male sex. Their man body mass index was 28.0 kg/m^2^ for male and 27.3 kg/m^2^ for female in the European ethnic population. The study included in the GIANT meta-analysis for body mass index had median age ranges from 40s to 70s. Their median body mass index among studies ranged from 21.1 to 35.5 in males and 20.4 to 37.9 in females.

### MR results in the UK Biobank

After Steiger filtering and harmonization process, 282, 292, 306, and 299 SNPs were used in the MR analysis towards body mass index, waist circumference, hip circumference and waist-to-hip ratio (Supplementary Table 1). The remaining SNPs had genome-wide significant association with tobacco smoking initiation but were weakly bond to outcome phenotypes, ensuring the direction of the MR analysis is from the genetically predicted tobacco use towards obesity-related traits.

The main causal estimates by multiplicative random-effect inverse variance weighted method provided consistently significant results from higher prevalence of tobacco smoking on higher body mass index [from two-fold increase in prevalence, +0.28 (0.18, 0.38) kg/m^2^], higher waist circumference [+0.88 (0.66, 1.10) cm], higher hip circumference [+0.40 (0.23, 0.57)], and higher waist-to-hip ratio [+0.006 (+0.005, 0.007)] (Figure 2 and Table 2).

**Figure 2.**
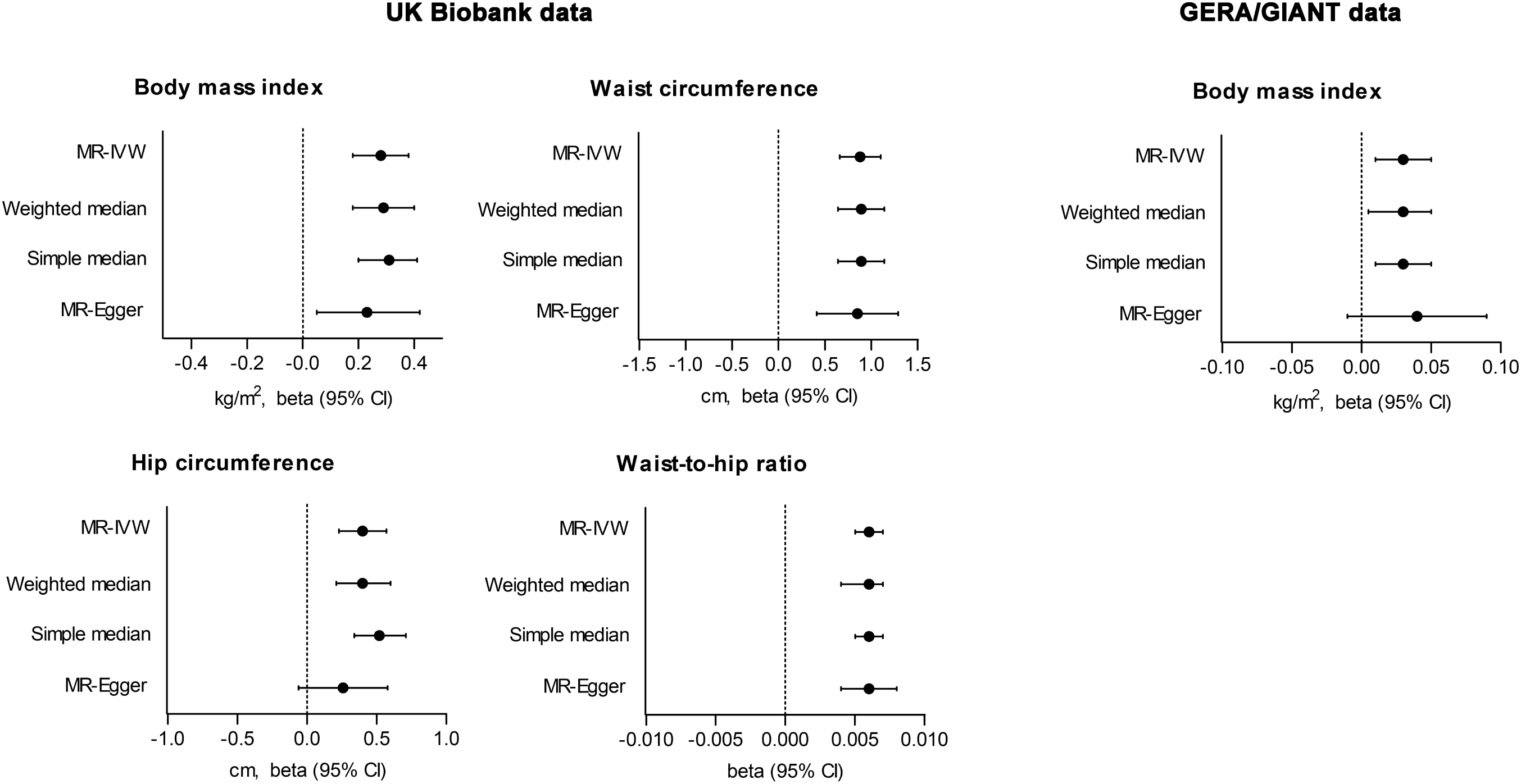
Causal estimates from the Mendelian randomization analysis. The x-axes show the causal estimates. The esimates by MR-IVW (multiplicative random-effect inverse variance weighted method), simple median, weighted median, and MR-Egger regression with bootstrapped standard error are presented. The outcome phenotpyes assessed in the UK Biobank were body mass index, waist circumference, hip circumference, and waist-to-hip ratio. The replication analysis was performed for GERA/GIANT consortium data towards body mass index outcome. Causal estimates are scaled towards 2-fold increase in prevalence of tobacco smoking.

**Table 2.** Causal estimates in the UK Biobank data.

| Outcome | N of instrumented SNPs | MR-Egger intercept P value | MR methods | Causal estimates [beta (95% CI)] | P value |
| --- | --- | --- | --- | --- | --- |
| Body mass index (kg/m<sup>2</sup>) | 282 | 0.109 | MR-IVW | 0.28 (0.18, 0.38) | < 0.001 |
|  |  |  | Weighted median | 0.29 (0.18, 0.4) | < 0.001 |
|  |  |  | Simple median | 0.31 (0.2, 0.41) | < 0.001 |
|  |  |  | MR-Egger | 0.23 (0.05, 0.42) | 0.005 |
| Waist circumference (cm) | 292 | 0.116 | MR-IVW | 0.88 (0.66, 1.1) | < 0.001 |
|  |  |  | Weighted median | 0.89 (0.64, 1.14) | < 0.001 |
|  |  |  | Simple median | 0.89 (0.64, 1.14) | < 0.001 |
|  |  |  | MR-Egger | 0.85 (0.41, 1.29) | < 0.001 |
| Hip circumference (cm) | 306 | 0.253 | MR-IVW | 0.4 (0.23, 0.57) | < 0.001 |
|  |  |  | Weighted median | 0.4 (0.21, 0.6) | < 0.001 |
|  |  |  | Simple median | 0.52 (0.34, 0.71) | < 0.001 |
|  |  |  | MR-Egger | 0.26 (-0.06, 0.58) | 0.06 |
| Waist-to-hip ratio | 299 | 0.523 | MR-IVW | 0.006 (0.005, 0.007) | < 0.001 |
|  |  |  | Weighted median | 0.006 (0.004, 0.007) | < 0.001 |
|  |  |  | Simple median | 0.006 (0.005, 0.007) | < 0.001 |
|  |  |  | MR-Egger | 0.006 (0.004, 0.008) | < 0.001 |

MR = Mendelian randomization, SNP = single nucleotide polymorphism, MR-IVW = multiplicative random-effect inverse-variance weighted method
Causal estimates are scaled towards 2-fold increase in prevalence of tobacco smoking.

The causal estimates remained significant towards all studied outcomes in the UK Biobank data by the median-based methods with the same directional effect. The results were even supported by MR-Egger regression with bootstrapped standard error with similar effect sizes towards body mass index [+0.23 (0.05, 0.42) kg/m^2^], waist circumference [+0.85 (0.41, 1.29) cm], and waist-to-hip ratio [+0.006 (0.004, 0.007)]. The result towards hip circumference was attenuated [+0.26 (−0.06, 0.58) cm], however, MR-Egger intercept P value indicated low probability of directional pleiotropy throughout the analysis (intercept P > 0.05).

The results were similarly identified in our sensitivity analysis after disregarding the SNPs that were identified to have notable association with other traits in the GWAS catalog (Supplementary Table 3), and genetically predicted tobacco smoking was consistently associated with worse obesity-related traits.

### MR results in the GERA/GIANT data

When applying the Stieiger-filtered SNPs towards the GERA/GIANT data, 275 SNPs were utilized to genetically predict tobacco smoking initiation (Supplementary Table 2). The MR analysis demonstrated that genetically predicted tobacco smoking was significantly associated with higher body mass index [+0.03 (0.01, 0.05)], supported by the weighted median [+0.03 (0.00, 0.05)] and the simple median [+0.03 (0.01, 0.05)] methods (Figure 2 and Table 3). MR-Egger regression with bootstrapped error also yielded significant causal estimates [+0.04 (0.00, 0.08)], and MR-Egger intercept P value indicated low probability of a directional pleiotropy (intercept P = 0.40). The results were similarly reproduced when the SNPs with potential association with other traits screened in the GWAS catalog (Supplementary Table 4)

**Table 3.** Causal estimates in the GERA/GIANT meta-analysis.

| Outcome | N of instrumented SNPs | MR-Egger intercept P value | MR methods | Causal estimates [beta (95% CI)] | P value |
| --- | --- | --- | --- | --- | --- |
| Body mass index (kg/m<sup>2</sup>) | 275 | 0.402 | MR-IVW | 0.03 (0.01, 0.05) | 0.01 |
|  |  |  | Weighted median | 0.03 (0, 0.05) | 0.03 |
|  |  |  | Simple median | 0.03 (0.01, 0.05) | 0.02 |
|  |  |  | MR-Egger | 0.04 (-0.01, 0.09) | 0.045 |

MR = Mendelian randomization, SNP = single nucleotide polymorphism, MR-IVW = multiplicative random-effect inverse-variance weighted method
Causal estimates are scaled towards 2-fold increase in prevalence of tobacco smoking.

## Discussion

In this MR analysis, we identified that genetically predicted initiation of tobacco smoking was significantly associated with worse obesity-related traits. Considering the data characteristics, our study suggests that being an ever smoker, mostly initiated in adolescents or in the early adulthood, would causally lead to higher body mass index, higher waist circumference, and higher wait-to-hip ratio in the later life period. Our study supports that tobacco smoking may consequently lead to worse obesity-related traits and that the misbelief for weight control effect from tobacco smoking may not hold in long-term.

The causal linkage between obesity and tobacco use has been suggested before.^7^ The previous study reported that being obesity itself is a risk factor to initiate tobacco use. Obesity-related social disadvantages may be associated with the initiation of tobacco use in the population, and the findings supported the close association between obesity and tobacco use in a previous observational report.^25^ On the other hand, the causal effects from being an ever smoker in obesity-related traits in the middle-to-old age population have been yet determined by MR analysis, which is a useful tool to investigate causal effects from modifiable exposure on complex outcomes. A previous study addressed that high tobacco consumption may lower weight by MR analysis,^26^ however, the message is limited to current smokers and a single-SNP analysis is easy to be affected from horizontal pleiotropic effect and weak instrumental bias.^27^ Our study has strengths in that 1) we used a strong genetic instrumental variable towards being an ever smoker based from previous large-scale GWAS meta-analysis, 2) we paid particular attention to the reverse directional genetic effect by Setiger filtering, 3) potential confounder effects by pleiotropy-robust sensitivity analyses, 4) and replicated the findings in another independent outcome summary-level data. With the consistent results, our MR results supported that being an ever smoker would be a causal risk factor for obesity.

The notable point is that the smoking initiation mostly occurred in adolescents or in young adulthoods and the obesity-related traits were measured in middle-to-old age individuals. Thus, this study suggests that smoking initiation in early period of life would consequently lead to increased body weight in long-term. The evidence is important to educate the young population not to start tobacco use, particularly affected from the misbeliefs that tobacco smoking may have weight control effect.^3,4^ Although the short-term appetite suppression effect from nicotine would be present,^5^ tobacco smoking may lead to unhealthy behaviors or social factors contributing to obesity in long-term.^28,29^ In addition, having no history of tobacco smoking would be helpful for physical activity and other healthy lifestyles, supporting the identified causal effects from being an ever smoker on higher body mass index and waist circumference.^30^ Thus, being non-smoker would be the best way to lower the risk of obesity in later period life and people should not initiate tobacco use in anticipation for long-term weight control effect, which also elevates the risk of diabetes and metabolic syndrome.^31,32^ In addition, considering that obesity is one of the most important risk factors for cardiovascular diseases and mortality in modern medicine, policies should also consider reduction in tobacco initiation rate in the population would finally ameliorate the substantial socioeconomic burden related to obesity.

There are limitations in the study. First, the outcome data had some overlap with the dataset provided the genetic instrumental variable.^16^ However, in the replication analysis, the proportion was small and considering the previous simulation study, the potential bias would not substantially influence the findings. Second, performing MR analysis with binary exposure tests the association between genetic liability and outcomes.^23^ However, utilizing quantitative traits (e.g. smoking heaviness) as exposure phenotypes was not possible for tobacco smoking as testing the causal estimates only in a subgroup (e.g. smokers) causes collider bias, thus the effect within smokers would require different study design.^33^ Third, MR analysis cannot determine the mediating mechanism from being an ever smoker on higher body mass index. There may be various vertical pathways (e.g. healthy lifestyles related to being a non-smoker or social factors in tobacco users) which warrants additional study. Lastly, the study was performed mostly within European ancestry individuals, thus the generalizability to other ethnic population should be expanded in the future.

In conclusion, genetically predicted being an ever smoker causally leads to worse obesity-related traits in middle-to-old age populations. Young people should be educated that initiation of tobacco use would consequently lead to higher risk of obesity in the later periods of life. Healthcare providers and policy makers should mind the adverse effect from tobacco initiation on the burden of obesity and move forward to prevent the people from starting tobacco use.

## Data Availability

The data used for this study is available in the public database. The CKDGen data is openly available in the consortium website (URL: https://ckdgen.imbi.uni-freiburg.de/) and the UK Biobank data is accessible after acquiring approval from the organization (application No. 53799).

## Authors’ contribution

The corresponding author attests that all listed authors meet the authorship criteria and that no others meeting the criteria have been omitted. SP, HL, KSK, KWJ, and DKK contributed to the conception and design of the study. SL, SK, YK, YCK, SSH, JPL, KWJ, CSL, YSK, and DKK provided statistical advice and interpreted the data. SP, KSK performed the main statistical analysis, assisted by SK, SL and YK. HL, JPL, KWJ, CSL, YSK, and DKK provided advice regarding the data interpretation. YCK, SSH, HL, JPL, KWJ, CSL, and YSK provided material support during the study. SP and DKK had full access to all data in the study and take responsibility for the integrity of the data and the accuracy of the data analysis. All authors participated in drafting the manuscript. All authors reviewed the manuscript and approved the final version to be published.

## Acknowledgements

The study was based on the data provided by the UK Biobank consortium (application No. 53799). We thank the investigators of the CKDGen consortium who provided the summary statistics for the outcome data of this study.

