## Supplementary Material for "Causal effects from tobacco smoking on obesity-related traits: a Mendelian randomization study"

S Park et al.

**Table of contents**

**Supplementary Table 1**. Information of genetic instruments and their association with outcome phenotypes in the UK Biobank data.

**Supplementary Table 2**. Information of genetic instruments and their association with outcome phenotypes in the GERA/GIANT data.

**Supplelmentary Table 3.** Causal estimates in the UK Biobank data by sensitivity analysis with trimmed genetic instrumental variable.

**Supplementary Table 4**. Causal estimates in the GERA/GIANT meta-analysis by sensitivity analysis with trimmed genetic instrumental variable.

**Supplelmentary Table 3.** Causal estimates in the UK Biobank data by sensitivity analysis with trimmed genetic instrumental variable.

| **Outcome** | **N of instrumented SNPs** | **MR-Egger intercept P value** | **MR methods** | **Causal estimates [beta (95% CI)** | **P value** |
| --- | --- | --- | --- | --- | --- |
| Body mass index (kg/m^2^) | 274 | 0.09 | MR-IVW | 0.28 (0.18, 0.38) | < 0.001 |
|  |  |  | Weighted median | 0.29 (0.18, 0.4) | < 0.001 |
|  |  |  | Simple median | 0.31 (0.2, 0.41) | < 0.001 |
|  |  |  | MR-Egger | 0.25 (0.07, 0.43) | 0.001 |
| Waist circumeference (cm) | 281 | 0.11 | MR-IVW | 0.86 (0.63, 1.08) | < 0.001 |
|  |  |  | Weighted median | 0.87 (0.62, 1.12) | < 0.001 |
|  |  |  | Simple median | 0.87 (0.62, 1.12) | < 0.001 |
|  |  |  | MR-Egger | 0.83 (0.39, 1.27) | < 0.001 |
| Hip circumference (cm) | 295 | 0.24 | MR-IVW | 0.4 (0.22, 0.57) | < 0.001 |
|  |  |  | Weighted median | 0.42 (0.23, 0.61) | < 0.001 |
|  |  |  | Simple median | 0.54 (0.34, 0.73) | < 0.001 |
|  |  |  | MR-Egger | 0.26 (-0.09, 0.61) | 0.07 |
| Waist-to-hip ratio | 290 | 0.35 | MR-IVW | 0.006 (0.005, 0.007) | < 0.001 |
|  |  |  | Weighted median | 0.006 (0.004, 0.007) | < 0.001 |
|  |  |  | Simple median | 0.006 (0.005, 0.008) | < 0.001 |
|  |  |  | MR-Egger | 0.006 (0.004, 0.009) | < 0.001 |

MR = Mendelian randomization, SNP = single nucleutide polymorphism, MR-IVW = multiplicative random-effect inverse-variance weighted method

Causal estimates are scaled towards 2-fold increase in prevalence of tobacco smoking.

**Supplementary Table 4**. Causal estimates in the GERA/GIANT meta-analysis by sensitivity analysis with trimmed genetic instrumental variable.

| **Outcome** | **N of instrumented SNPs** | **MR-Egger intercept P value** | **MR methods** | **Causal estimates [beta (95% CI)** | **P value** |
| --- | --- | --- | --- | --- | --- |
| Body mass index (kg/m^2^) | 267 | 0.33 | MR-IVW | 0.03 (0.01, 0.06) | 0.008 |
|  |  |  | Weighted median | 0.03 (0.01, 0.05) | 0.02 |
|  |  |  | Simple median | 0.03 (0.01, 0.05) | 0.01 |
|  |  |  | MR-Egger | 0.04 (0.00, 0.09) | 0.049 |

MR = Mendelian randomization, SNP = single nucleutide polymorphism, MR-IVW = multiplicative random-effect inverse-variance weighted method

Causal estimates are scaled towards 2-fold increase in prevalence of tobacco smoking.
